# Explainable Deep Learning Reveals Distributed Neurodegeneration Signatures of Neuropsychiatric Symptoms Across the Alzheimer’s Continuum

**DOI:** 10.64898/2026.08.24.26361256

**Authors:** Bahram Yaghooti, Saba Ishrat, Hoang Nam Le, Ram P. Sapkota, Taslim Murad, Deepa S. Thakuri, Dean F. Wong, Andrew Aschenbrenner, J. Philip Miller, Justin M. Long, Ginger E. Nicol, Eric J. Lenze, Alzheimer’s Disease Neuroimaging Initiative, Ganesh B. Chand

**Affiliations:** Neuroimaging Informatics and Artificial Intelligence (NeuroAI) Laboratory, Department of Radiology, Mallinckrodt Institute of Radiology, Washington University School of Medicine, St. Louis, MO, USA; Department of Computer Science, Institute of Business Administration, Karachi, Pakistan; School of Medicine, University of Missouri, Columbia, MO, USA; Departments of Neurosurgery, Radiology and Psychiatry, Kahlert Institute of Addiction Medicine, University of Maryland Baltimore School of Medicine, Baltimore, MD, USA; Department of Neurology, University of Kansas, Kansas City, KS, USA; Institute for Informatics, Data Science and Biostatistics, Washington University School of Medicine, St. Louis, MO, USA; Department of Neurology, Washington University School of Medicine, St. Louis, MO, USA; Knight Alzheimer Disease Research Center, Washington University School of Medicine, St. Louis, MO, USA; Department of Psychiatry, Washington University School of Medicine, St. Louis, MO, USA; NeuroGenomics and Informatics Center, Washington University School of Medicine, St. Louis, MO, USA; Roy and Diana Vagelos Division of Biology & Biomedical Sciences – Neurosciences; Biomedical Informatics and Data Science; Computational and Systems Biology; Biochemistry, Biophysics, and Structural Biology, Washington University School of Medicine, St. Louis, MO, USA

**Author notes:** **Corresponding authors:** Bahram Yaghooti, PhD and Ganesh B. Chand, PhD. **Funding:** NIH/NIA RF1AG087271.

**Keywords:** Alzheimer’s disease, neuropsychiatric symptoms, NPI-Q, structural MRI, explainable artificial intelligence, deep learning, SHAP, neurodegeneration, brain morphometry

## Abstract

Neuropsychiatric symptoms (NPS) are increasingly recognized as critical components of the disease progression in Alzheimer’s disease (AD), yet their relationship with neurodegeneration remain poorly characterized. We investigated the multivariate relationships between structural MRI (sMRI)-based regional neurodegenerative biomarkers and NPS using the Alzheimer’s Disease Neuroimaging Initiative (ADNI) and Knight Alzheimer Disease Research Center (Knight-ADRC) cohorts (*N* = 1,756). The machine learning regression models were compared for NPS prediction, and the best-performing deep neural network (named NPSNet) was integrated with three feature-importance methods: SHapley Additive exPlanations (SHAP), Local Interpretable Model-agnostic Explanations (LIME), and Layer-wise Relevance Propagation (LRP). To establish a known ground truth, we introduced predefined regional perturbations into semi-simulated data and tested whether NPSNetSHAP, NPSNetLIME, and NPSNetLRP could recover them. The NPSNet strongly predicted NPS scores (pooled Spearman *ρ* = 0.927, *p* < 2.2 × 10⁻³⁰⁸; fold-wise *ρ* = 0.912 − 0.963) and NPSNetSHAP recovered all 100% perturbed regions, compared with 90% for NPSNetLIME and 40% for NPSNetLRP. In the experimental data (*N* = 1,756), the NPSNet produced the highest held-out correlation (*ρ* = 0.387, *p* = 1.7 × 10⁻¹⁵), exceeding gradient boosting (*ρ* = 0.301), support vector regression (*ρ* = 0.266), and others (*ρ* < 0.266). NPSNetSHAP identified individual-level regional contribution patterns relevant to NPS predictions. Comparing NPSNetSHAP attributions between cognitively normal (CN) and mild cognitive impairment (MCI)/AD groups revealed distributed multivariate neurodegenerative signatures of NPS, with the largest differences between CN and AD participants. This study introduces an explainable deep learning framework for identifying distributed, individualized neurodegeneration signatures of NPS burden across the AD continuum.

## Introduction

Alzheimer’s disease (AD) is a progressive neurodegenerative disorder and the most common cause of dementia (1). Although AD is primarily characterized by progressive cognitive decline, neuropsychiatric symptoms (NPS), including apathy, depression, anxiety, irritability, agitation, disinhibition, sleep disturbances, delusions, and hallucinations, commonly occur during symptomatic stages of cognitive impairment and dementia. Some NPS may emerge during mild cognitive impairment (MCI), before dementia becomes clinically evident (2, 3). NPS are associated with increased patient distress, caregiver burden, and the likelihood of institutional care (4–6). In individuals with MCI, specific NPS profiles have been associated with a less favorable clinical course and a greater likelihood of conversion to dementia. For example, apathy- and irritability-dominant profiles have been associated with increased conversion risk, although conversion may be to dementia due to AD or another form of dementia (7). Thus, NPS may provide prognostic information about the clinical progression of MCI, but they do not by themselves indicate the presence or progression of biologically defined AD. The underlying neurodegenerative signatures of NPS remain incompletely understood.

The neuroanatomical basis of NPS in AD is still not fully understood. AD-related neurodegeneration affects multiple interacting brain systems involved in emotional regulation, motivation, perception, impulse control, and social behavior. Alterations involving limbic, frontal, temporal, parietal, and subcortical regions may therefore contribute jointly to an individual’s NPS profile (8, 9). Structural magnetic resonance imaging (sMRI) provides in vivo measures of regional brain morphology, allowing assessment of structural alterations across gray matter, white matter, and ventricular/cerebrospinal fluid regions collectively (10). Previous sMRI studies have associated NPS with atrophy in frontal, temporal, limbic, and association regions. Psychotic symptoms have been linked to neocortical atrophy, whereas agitation and aggression have been associated with alterations in frontolimbic and salience-network regions (11, 12). Cortical thinning has also been associated with worsening apathy and hallucinations (13).

Some of this variation may be related to the way these relationships have been studied (14–16). Many previous analyses have examined the relationships between NPS and individual brain regions, considering one region at a time (11, 17, 18). A whole-brain multivariate approach is particularly relevant because similar symptoms may arise from different structural patterns, and changes in the same region may have different behavioral effects depending on the condition of the rest of the brain. Among multivariate approaches, deep learning is well suited to this problem because it can learn nonlinear and higher-order interactions among numerous correlated regional features without requiring the form of those relationships to be specified in advance. Because this flexibility may also increase the risk of overfitting, the deep neural networks for NPS prediction (named NPSNet) model used in the present analysis was compared directly with regularized linear regression, support vector regression, and gradient boosting to determine whether it improved held-out prediction rather than assuming its superiority.

Recent work using whole-brain sMRI measures has also shown that associations between NPS and brain morphology may differ between MCI and AD dementia (15). However, that study included 235 participants limited to MCI and AD dementia, without a cognitively normal comparison group, and used region-wise linear association models. Although this approach provided whole-brain anatomical coverage, it was not designed to jointly model distributed regional features, capture nonlinear or higher-order interregional relationships, or generate individualized out-of-sample predictions. These capabilities are relevant because NPS contribute to clinical heterogeneity and burden in MCI and AD dementia, and generalizable multiregional models may help characterize distributed structural patterns associated with individual differences in NPS burden. Deep learning may therefore provide a flexible framework for modeling these relationships across clinically defined cognitive-status groups when its performance is supported by comparison with alternative multivariate models.

In the present study, NPS burden was assessed using the Neuropsychiatric Inventory Questionnaire (NPI-Q) (5). We selected the total NPI-Q severity score as the primary outcome because the study was designed to model overall NPS burden across the AD continuum rather than the anatomical substrate of any single symptom. The total score provided a common continuous outcome across both cohorts, preserved statistical power, and avoided fitting multiple comparatively sparse domain-specific models. It was also well suited to evaluating whether a distributed whole-brain patterns could capture the combined burden of clinically heterogeneous symptoms. Higher scores indicate greater overall symptom burden affecting patients and caregivers that is not captured by standard cognitive screening measures.

Deep learning methods can examine relationships among many correlated regional features simultaneously. Most ML studies in AD neuroimaging, however, have focused on diagnostic classification, cognitive prediction, or disease progression, with less attention given to predicting overall NPS burden from whole-brain neurodegenerative measures. Moreover, prediction performance alone does not reveal which brain regions contribute to a model’s output. Explainable machine learning /artificial intelligence (ML/AI) methods address this limitation, but they generate explanations using different assumptions and computational strategies. SHAP estimates additive feature contributions using principles from cooperative game theory (19), LIME approximates individual predictions by fitting interpretable surrogate models to locally perturbed observations (20), and LRP propagates relevance backward through the layers of a neural network (21). These methods therefore represent complementary game-theoretic, model-agnostic, and network-specific approaches and may produce substantially different regional rankings when applied to the same predictive model. Because the ground-truth regional contributions are unknown in observational neuroimaging data, an anatomically plausible explanation cannot by itself establish that an attribution method is accurate. We therefore compared SHAP, LIME, and LRP in semi-simulated data with predefined perturbed regions before applying and interpreting the experimental data. This validation provided an empirical basis for selecting the method that most faithfully recovered the known patterns and reduced the risk of interpreting method-dependent artifacts as biologically meaningful findings.

Herein, we investigated whether whole-brain neurodegenerative signatures derived from regional sMRI volumes could predict NPS scores across the AD continuum. We compared a NPSNet with gradient boosting, support vector regression, Lasso, Elastic Net, and Ridge regression models. The central methodological innovation was a two-stage explainable deep-learning framework that evaluated predictive performance and explanation fidelity separately before interpreting experimental data. First, the best-performing predictive model was identified through direct comparison with conventional linear and nonlinear methods. Second, SHAP, LIME, and LRP were applied to the same trained NPSNet and evaluated using semi-simulated data in which the perturbed brain regions were known. Only after an attribution method demonstrated that it could recover this ground-truth pattern was it used to identify individualized regional contributions in the experimental data and compare cognitively normal, MCI, and AD dementia diagnosis groups. This design provided an empirical validation step that is absent when post hoc explanations are applied directly to observational neuroimaging data. We hypothesized that the NPSNetSHAP would capture distributed multivariate relationships between whole-brain neurodegeneration signatures and NPS burden, that the selected explanation method would recover the simulated ground-truth regions, and that individualized regional contributions would differ across the AD continuum.

## Materials and Methods

### 2.1 Experimental data

Baseline T1-weighted sMRI data were obtained from the Alzheimer’s Disease Neuroimaging Initiative phase 2 (ADNI-2) (10) and the Knight Alzheimer Disease Research Center (Knight-ADRC) cohorts (22, 23). We included participants with an available baseline NPI-Q assessment who met the cohort-specific inclusion and exclusion criteria described in the **Supplementary Materials** (**Section S1**). When more than one NPI-Q assessment was available, the assessment obtained closest to the sMRI acquisition date was selected. The combined experimental dataset included 1,756 participants. The ADNI dataset included 668 participants, of whom 308 were female. Their ages ranged from 55.1 to 91.5 years, with a mean age of 72.8 years and a standard deviation of 7.3 years. Based on the diagnostic categories available in ADNI, 187 participants were cognitively normal (CN), 173 had early EMCI, 152 had late MCI (LMCI), 6 had MCI, and 150 had AD dementia. For the group analyses, participants with EMCI, LMCI, or MCI were combined into a single MCI group. The Knight-ADRC dataset included 1,088 participants, of whom 605 were female. Their ages ranged from 45.2 to 91.4 years, with a mean age of 70.4 years and a standard deviation of 8.5 years. This cohort included 920 CN participants and 168 participants in a combined symptomatic category. For the group analyses, Knight-ADRC participants were classified using global CDR-based criteria, and the symptomatic categories were combined because of the limited number of cognitively impaired participants. The predominance of CN participants reflects the Knight-ADRC’s research focus on cognitively unimpaired individuals and preclinical AD. The two cohorts were combined to provide a larger and more heterogeneous sample covering different ages and levels of cognitive impairment. The demographic and diagnostic characteristics of both cohorts are summarized in **Table 1** and detailed in the **Supplementary Materials** (**Section S1**).

**Table 1.** Summary of experimental data. Demographic and clinical cognitive-status characteristics of the ADNI (n = 668) and Knight-ADRC (n = 1,088) cohorts. Counts are shown for clinical cognitive status and sex; age is reported in years. Clinical cognitive-status categories were defined independently of amyloid biomarker status. Knight-ADRC participants with a global CDR score of 0.5 or higher were combined into a symptomatic group for analysis. The availability and distribution of amyloid biomarker data and global CDR scores are described in the **Supplementary Materials** (**Section S1**).

| Data Cohort | Diagnosis |  |  |  |  | Sex |  | Age (years) |  |  |  |
| --- | --- | --- | --- | --- | --- | --- | --- | --- | --- | --- | --- |
|  | CN | EMCI | LMCI | MCI | AD | Male | Female | Min | Max | Mean | Std |
| ADNI | 187 | 173 | 152 | 6 | 150 | 360 | 308 | 55.1 | 91.5 | 72.8 | 7.3 |
| Knight ADRC | 920 | 168 |  |  |  | 483 | 605 | 45.2 | 91.4 | 70.4 | 8.5 |

The NPS burden was measured using the NPI-Q total severity score (5). The NPI-Q is a brief informant-based assessment of behavioral and psychological symptoms observed during the preceding month. For each endorsed symptom, the informant rates its severity as mild, moderate, or severe and reports the associated caregiver distress. Severity ratings are summed across symptom domains to produce a total score ranging from 0 to 36, with higher scores indicating greater NPS burden. Caregiver-distress scores were not included in the present analyses. Each sMRI scan was segmented into 145 anatomical regions of interest using the Multi-atlas region Segmentation utilizing Ensembles of registration algorithms and parameters and Locally Optimal Atlas Selection (MUSE) method (24–27). These regions covered cortical and subcortical gray matter, white matter, and cerebrospinal fluid/ventricular structures, providing regional measures across the whole brain. The regional volumetric measures were harmonized using a previously developed data harmonization method to adjust for imaging site, cohort, age, sex, and total intracranial volume (25, 28, 29). The harmonized regional volumes were used as inputs to the ML/AI models, and the NPI-Q total severity score was used as the prediction target.

### 2.2 Semi-simulated data

A semi-simulated dataset was generated to determine whether the proposed NPSNetSHAP framework could superiorly recover a predefined pattern of multivariate associations between regional brain features and NPI-Q scores than other approaches. The simulation was based on the MUSE-derived and harmonized regional imaging features from the combined ADNI and Knight-ADRC cohorts. The ten regions showing the strongest absolute correlations with the NPI-Q outcome among the 145 regional features were selected for perturbation. These regions were the bilateral hippocampus, bilateral amygdala, bilateral entorhinal area, bilateral precuneus, right hippocampal region, and right parahippocampal gyrus. The semi-simulated analysis used the 10 selected regions to create the predefined ground-truth perturbation pattern. The full set of 145 regional features was used subsequently for modelling the experimental dataset. This data-driven selection allowed the attribution methods to be evaluated according to their ability to recover the regions with imposed perturbations. The final semi-simulated dataset comprised 2,000 participants generated from the 1,107 control participants using different participant-specific perturbations. Of these, 1,000 participants (50%) were assigned to the perturbed group, while the remaining 1,000 served as the unperturbed comparison group. For perturbed participants, the volumes of the selected regions were reduced while the corresponding NPI-Q scores were increased. This procedure imposed a known inverse relationship between regional brain integrity and NPS burden while retaining the natural variability and covariance structure of the imaging data. The perturbed regions therefore served as the ground truth against which the feature-attribution methods could be evaluated. The resulting dataset was used both to assess NPI-Q prediction and to compare the ability of SHAP, LIME, and LRP to recover the predefined regions among their most influential imaging features. The complete generation process for the semi-simulated data is described in the **Supplementary Materials** (**Section S2**). Cross-validated prediction performance on semi-simulated data is detailed in the **Supplementary Materials** (**Section S3**)

### 2.3 NPI-Q prediction using machine and deep learning models

We evaluated six supervised regression models for predicting the NPI-Q total score from the harmonized and normalized regional sMRI features. These modelling methods (30–34) included Lasso regression, Ridge regression, Elastic Net, support vector regression (SVR), gradient boosting, and deep neural network for NPS prediction (named NPSNet). The regularized linear models were included to provide interpretable baselines and to address collinearity among regional brain measures. SVR and gradient boosting allowed nonlinear relationships to be modelled, whereas the NPSNet was used to capture more complex multivariate associations distributed across the brain. Hyperparameters for each conventional ML model were optimized through grid search using only the training data. Model selection was based on prediction performance within the training procedure, with mean absolute error used as the optimization criterion. The complete hyperparameter search spaces and the selected configurations is reported in the **Supplementary Materials** (**Section S4**). The NPSNet contained four fully connected hidden blocks with progressively fewer units: 40, 30, 20, and 10. This architecture was selected by comparing four candidate configurations in the semi-simulated data and produced the highest Spearman correlation (ρ = 0.796) and lowest mean absolute error (MAE = 0.880). The progressively narrowing design allowed the network to compress information from the 145 correlated regional inputs into increasingly compact representations while limiting model complexity and the risk of overfitting. Each block consisted of a dense layer with rectified linear unit activation, followed by batch normalization and dropout (35, 36). Batch normalization was used to stabilize training, while dropout provided additional regularization by randomly deactivating a proportion of the hidden units during each training iteration. The output layer contained a single unit with linear activation for predicting the continuous NPI-Q total score. The network weights were initialized using a random normal distribution. Training was monitored using an internal validation set, and early stopping was applied when the validation loss no longer improved. The weights associated with the lowest validation loss were restored before evaluation on the independent test fold. The NPSNet was implemented in Python using PyTorch (37). The learning rate, batch size, number of training epochs, dropout rate, and other network hyperparameters were selected through the tuning procedure described in the **Supplementary Materials** (**Section S4**).

### 2.4 NPSNetSHAP framework

The model demonstrating the strongest held-out NPI-Q prediction performance was carried forward for explainability analysis. Because the NPSNet outperformed the conventional ML models, it was selected as the best predictive model for downstream analyses. SHAP, LIME, and LRP were then compared using the semi-simulated data, in which the ten perturbed regions provided a known ground truth. NPSNetSHAP recovered all ten perturbed regions among its ten highest-ranked features, compared with nine for NPSNetLIME and four for NPSNetLRP. Based on this superior ground-truth recovery, SHAP was selected as the explanation method and integrated with the NPSNet to form the NPSNetSHAP framework for analysis of the experimental data.

SHAP quantifies the significance of an input feature for a model’s output through its SHAP value, which is derived from the Shapley value in cooperative game theory. Through Shapley value players in a game are assigned fair values based on their contribution to the game’s output. The SHAP value for an input feature (*i*) is computed through the given formula:

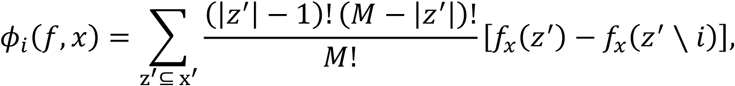

where, *f* denotes the trained model, (*x* = [*x*_1_, *x*_2_, …, *x*_*M*]) represents the input features, and *x*′ refers to simplified binary input vectors indicating which features are included (1) or excluded (0). Each subset *z*′ of *x*′ reflects a specific combination of present or absent features, and the term *f_x_*(*z*′) is the expected model output when only the features in *z*′ are known. The difference *f_x_*(*z*′) − *f_x_*(*z*′ ∖ *i*) captures the marginal effect of including feature *i*, and the weighting factor 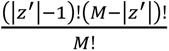 ensures fair contribution from each subset, derived from cooperative game theory principles.

A positive SHAP value indicates that a regional feature shifts the prediction toward a higher NPI-Q score relative to the model’s expected output, whereas a negative value shifts the prediction toward a lower score. The magnitude of the value reflects the strength of that contribution. For global interpretation, regional importance was calculated by averaging the absolute SHAP values across participants within each diagnostic groups. Regions with larger mean absolute values were therefore considered more influential to NPI-Q prediction. Individual SHAP values were retained for analyses examining participant-level and diagnostic-group patterns.

The semi-simulated dataset was also used to compare SHAP with LIME and LRP (20, 21). Each method was integrated with the trained NPSNet to produce NPSNetSHAP, NPSNetLIME, and NPSNetLRP explanations. Because the ten perturbed brain regions were known in advance, they provided a ground truth for evaluating the attribution methods. Performance was assessed according to the number of perturbed regions recovered among the ten highest-ranked features produced by each method. This comparison was used to determine which explanation method most faithfully recovered the imposed multivariate imaging pattern.

All predictive models were trained and evaluated using ten-fold cross-validation (38). In each fold, 90% of the observations were used for model development and the remaining 10% were held out for testing. Training and test observations remained separate within every fold. To reduce sensitivity to random initialization and data partitioning, the complete cross-validation procedure was repeated five times, and performance was averaged across the independent runs.

Predictive performance was assessed using Spearman’s rank correlation between the observed and predicted NPI-Q scores in the held-out test data. The corresponding p-value was used to evaluate whether the observed association differed significantly from zero. Brain maps were generated by projecting the regional attribution values onto a standard sMRI template and displaying them in the axial orientation. A detailed explanation of the explainable AI methods used in this paper is provided in the **Supplementary Materials** (**Section S5**).

### 2.5 Clinical severity under the NPSNetSHAP framework

To determine whether the imaging contributions identified by NPSNetSHAP varied across the Alzheimer disease continuum, individualized regional SHAP values were summarized separately for cognitively normal, MCI, and AD dementia groups. Participants with early MCI, late MCI, or an unspecified MCI diagnosis were included in the MCI group. A combined patient group containing participants with either MCI or AD was also examined to permit analysis across the diagnostic structure of the combined ADNI and Knight-ADRC dataset.

Group differences in the individualized NPSNetSHAP values were quantified using Cohen’s d (39). Comparisons were performed between cognitively normal participants and those with MCI, AD, or MCI/AD combined. These analyses were intended to identify regions whose contribution to NPI-Q prediction changed with increasing clinical severity. Because SHAP values describe a model’s use of a feature, rather than the direction of anatomical change itself, the resulting effect-size maps were interpreted as differences in regional predictive contributions and not as direct measures of atrophy or causal effects.

## Results

### 3.1 Semi-simulated data results

This section presents the performance of the conventional ML models and the NPSNet model for NPI-Q prediction using the semi-simulated dataset. **Table 2** summarizes the corresponding results, which indicate that the NPSNet model achieved superior performance compared with the conventional ML models.

**Table 2.** The performance of the conventional ML models and the NPSNet model for NPI-Q prediction.

| Rank | Model | Spearman $\rho$ | p-value |
| --- | --- | --- | --- |
| 1 | NPSNet | 0.927 | $< 2.2 \times 10^{-308}$ |
| 2 | Gradient boosting | 0.891 | $< 2.2 \times 10^{-308}$ |
| 3 | SVR | 0.876 | $< 2.2 \times 10^{-308}$ |
| 4 | Lasso | 0.842 | $< 2.2 \times 10^{-308}$ |
| 5 | Elastic Net | 0.835 | $< 2.2 \times 10^{-308}$ |
| 6 | Ridge | 0.823 | $< 2.2 \times 10^{-308}$ |

The NPSNet most accurately predicted NPI-Q scores in the semi-simulated dataset. Predicted and actual scores showed a strong positive Spearman correlation (*ρ* = 0.927, *p* < 2.2 × 10⁻³⁰⁸; **Figure 1**). The predictions generally followed the observed values across the range of NPI-Q scores, demonstrating that the model captured the multivariate relationships introduced between the regional imaging features and NPS burden.

**Figure 1.**
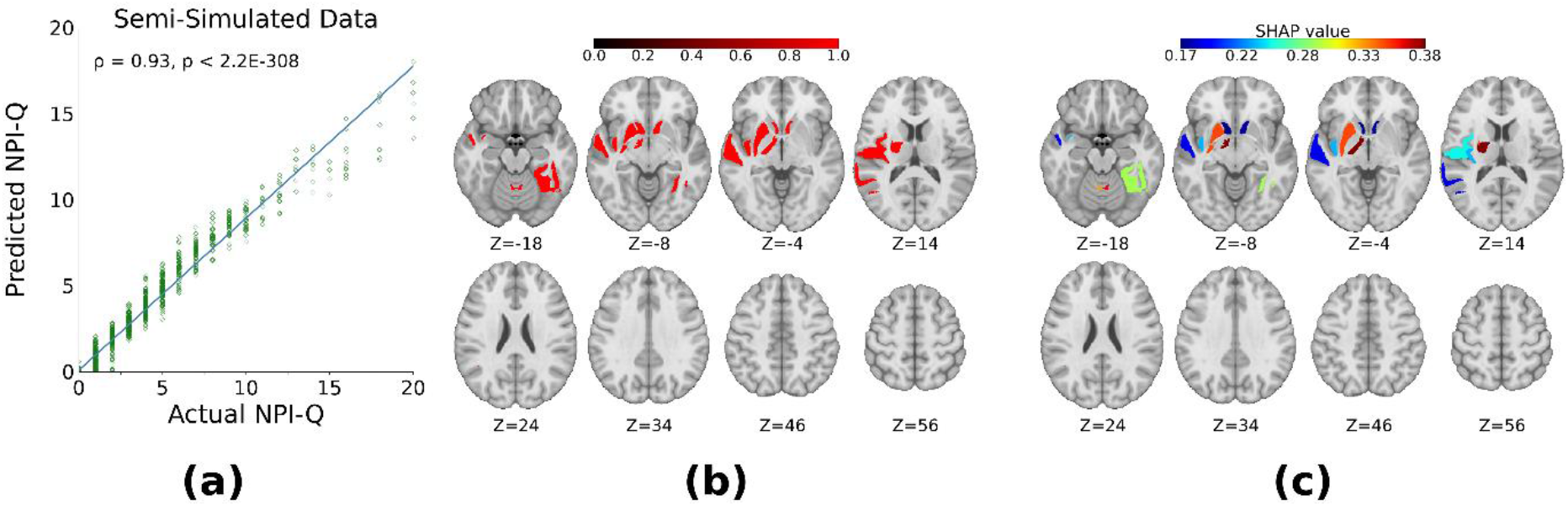
**(a)** Relationship between actual and NPSNet predicted NPI-Q scores in the semi-simulated dataset. The Spearman correlation coefficient and corresponding p-value are shown within the panel; **(b)** Top 10 regions chosen for perturbation; **(c)** Regions identified as significant using NPSNetSHAP framework.

The semi-simulated data also provided a known ground truth for evaluating the three feature-attribution methods (**Figure 2**). NPSNetSHAP recovered all ten perturbed regions among its ten highest-ranked features, corresponding to a recovery rate of 100%. These regions included the bilateral hippocampus, bilateral amygdala, bilateral entorhinal area, bilateral precuneus, right hippocampal and right parahippocampal gyrus. NPSNetLIME identified nine of the ten ground-truth regions, missing the right parahippocampal gyrus and yielding a recovery rate of 90%. In contrast, NPSNetLRP recovered only four of the ten perturbed regions: the bilateral amygdala, left entorhinal area and left precuneus. Its remaining highly ranked features were regions that had not been perturbed, resulting in a recovery rate of 40%. Thus, NPSNetSHAP demonstrated the closest agreement with the known regional patterns and was selected for interpreting NPI-Q predictions in the experimental dataset below.

**Figure 2.**
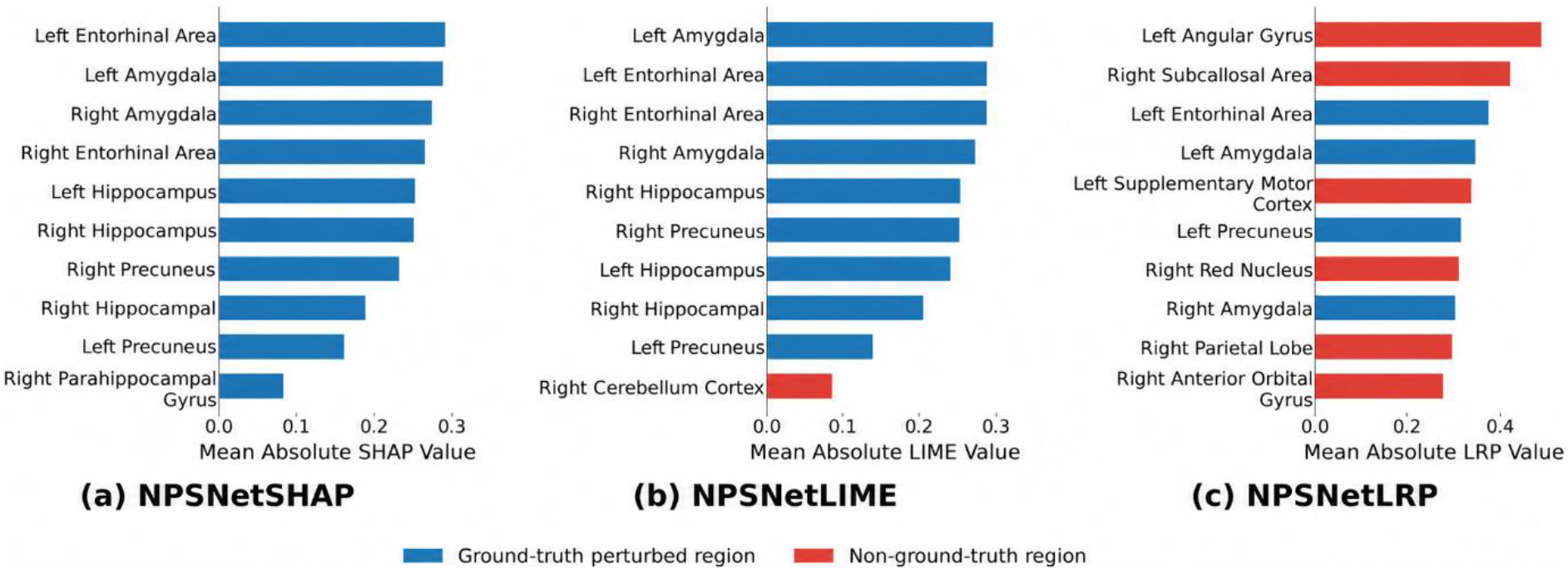
The ten regions with the largest mean absolute feature-attribution values for NPSNet predictions of NPI-Q total severity in the semi-simulated dataset, as estimated using **(a)** SHAP, **(b)** LIME, and **(c)** LRP. Blue bars denote regions included in the predefined ground-truth perturbation pattern; red bars denote unperturbed regions. “Influence” represents the magnitude of each region’s contribution to the model prediction and should not be interpreted as the direction of association or as evidence of an association in the experimental data.

### 3.2 Experimental data results

All six models produced significant positive correlations between observed and predicted NPI-Q scores in the combined ADNI and Knight-ADRC dataset (**Table 3**). The NPSNet achieved the strongest prediction performance, with a Spearman correlation of *ρ* = 0.387 (*p* = 1.7 × 10⁻¹⁵). Gradient boosting ranked second (*ρ* = 0.301, *p* = 4.1 × 10⁻¹³), followed by SVR (*ρ* = 0.266, *p* = 4.6 × 10⁻¹⁰). The regularized linear models showed weaker, although still significant, associations. Lasso produced a correlation of *ρ* = 0.214 (*p* = 6.6 × 10⁻⁷), while Elastic Net and Ridge achieved correlations of *ρ* = 0.205 (*p* = 2.0 × 10⁻⁶) and *ρ* = 0.202 (*p* = 2.8 × 10⁻⁶), respectively. The three nonlinear models occupied the highest positions in the performance ranking and these results showed that the multivariate relationships between regional whole-brain neurodegeneration measures and NPS burden was not fully captured by linear combinations of the imaging features. Because the NPSNet produced the highest held-out correlation, it was selected for the subsequent SHAP analysis.

**Table 3.** Performance of the ML/AI models for NPI-Q prediction in the experimental dataset.

| Rank | Model | Spearman $\rho$ | p-value |
| --- | --- | --- | --- |
| 1 | NPSNet | 0.387 | $1.7 \times 10^{-15}$ |
| 2 | Gradient boosting | 0.301 | $4.1 \times 10^{-13}$ |
| 3 | SVR | 0.266 | $4.6 \times 10^{-10}$ |
| 4 | Lasso | 0.214 | $6.6 \times 10^{-7}$ |
| 5 | Elastic Net | 0.205 | $2.0 \times 10^{-6}$ |
| 6 | Ridge | 0.202 | $2.8 \times 10^{-6}$ |

### 3.3 Regional NPSNetSHAP patterns across diagnostic groups

The NPSNetSHAP analysis revealed a spatially distributed pattern of regional contributions to NPI-Q prediction (**Figure 3(a)-(b)**). The mean absolute SHAP maps showed that the model drew on information from cortical, subcortical and ventricular regions rather than relying on a single anatomical structure. Contributions were visible across inferior and medial temporal regions, lateral temporal cortex, frontal and parietal areas, and structures surrounding the ventricular system. The cognitively normal and MCI groups showed relatively modest regional attribution magnitudes (**Supplementary Materials** (**Section S6**)). In contrast, the AD group displayed stronger and more spatially extensive NPSNetSHAP values, particularly across temporal and limbic regions. Elevated contributions were also evident in ventricular and periventricular regions. The combined MCI and AD group retained this broadly distributed pattern, although its mean attribution profile was less pronounced than that observed in the AD group alone. When all participants were considered together, the regional attribution map continued to show contributions distributed across multiple brain systems beyond the medial temporal regions most commonly associated in AD. Differences in individualized regional NPSNetSHAP values were examined across the diagnostic continuum using Cohen’s d effect sizes (**Figure 3(c); Supplementary Materials** (**Section S7**)).

**Figure 3.**
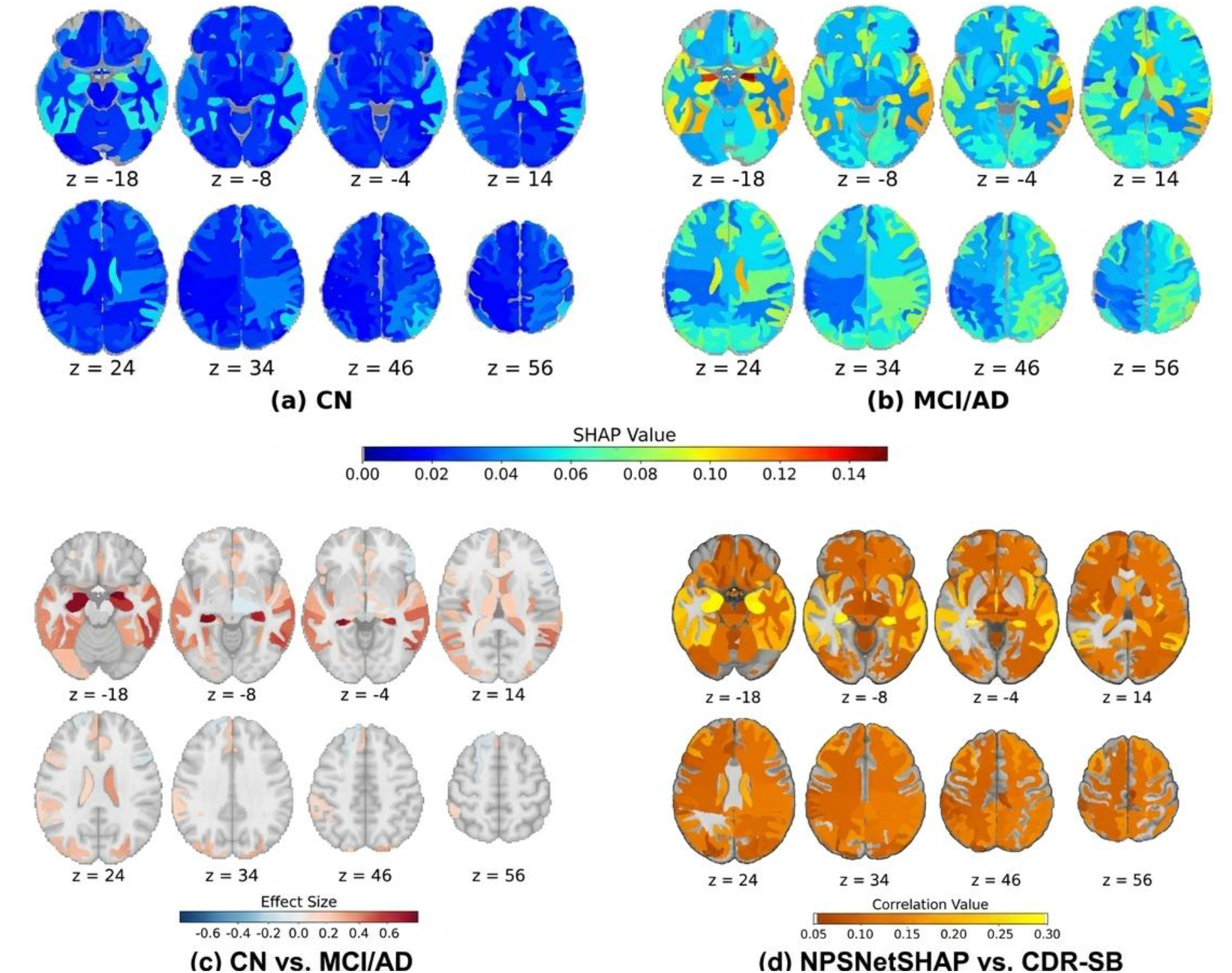
Regional NPSNetSHAP patterns associated with NPI-Q prediction and CDR-SB in the experimental dataset. Mean absolute SHAP values are displayed for (a) cognitively normal participants (CN) and (b) MCI/AD patients. Larger values indicate a stronger contribution of the corresponding regional imaging feature to the NPSNet prediction. Results are displayed on a standard sMRI template in axial orientation. (c) Cohen’s d effect size maps are shown for the comparisons between CN and MCI/AD Patients). (d) Spearman’s correlations between the individualized NPSNetSHAP regional features and CDR-SB scores across the AD continuum (CN, MCI, AD). The color bar highlights the corresponding values and z shows the brain slice position.

The relevance of individualized multivariate regional DL-SHAP features with clinical severity of AD was investigated by computing the Spearman’s correlations of these features with individuals’ CDR-SB score across the combined cohorts, and only the statistically significant correlations were considered. As demonstrated in **Figure 3(d)**, the brain regions with the strongest correlations with CDR-SB were the right amygdala, left hippocampus, left inferior lateral ventricle, right hippocampus, and right inferior lateral ventricle, followed by the bilateral entorhinal area, left amygdala, bilateral posterior insula, and bilateral middle temporal gyrus.

### 3.4 Regional NPSNetSHAP patterns according to diagnostic and NPI-Q status

To examine whether regional model contributions differed according to diagnosis and neuropsychiatric symptom status, participants were divided into four groups: cognitively normal participants with an NPI-Q score of zero, cognitively normal participants with a positive NPI-Q score, MCI/AD patients with an NPI-Q score of zero, and MCI/AD patients with a positive NPI-Q score. Mean absolute regional NPSNetSHAP values were calculated separately for each group and projected onto a standard sMRI template. The same color scale was used for all maps to allow direct visual comparison. The results are shown in **Figure 4**.

**Figure 4.**
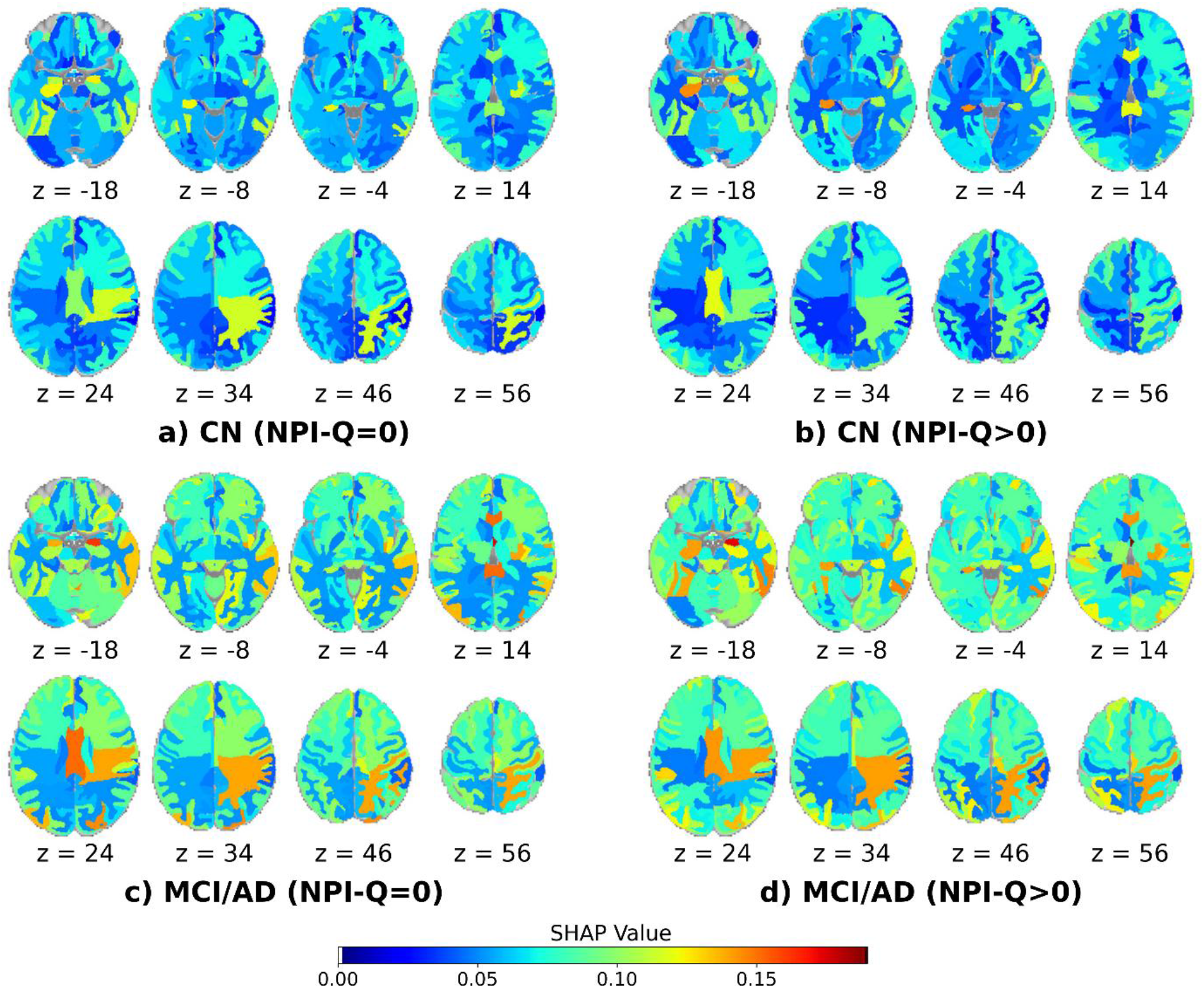
Regional NPSNetSHAP patterns according to diagnostic and NPI-Q status. Mean absolute SHAP values are shown for **(a)** cognitively normal participants with NPI-Q scores of zero [781 CN (NPI-Q=0)], **(b)** cognitively normal participants with positive NPI-Q scores [326 CN (NPI-Q>0)], **(c)** patients with NPI-Q scores of zero [194 MCI/AD (NPI-Q=0)], and **(d)** patients with positive NPI-Q scores [455 MCI/AD (NPI-Q>0)].

Among cognitively normal participants, both NPI-Q groups showed broadly distributed regional contributions to the model’s predicted NPI-Q total severity. Larger absolute SHAP values indicate greater contributions to the model prediction relative to its expected prediction. The overall spatial patterns were similar, although participants with positive NPI-Q scores showed focal increases in mean absolute SHAP values in several inferior and medial temporal regions, mainly involving the hippocampus, amygdala, entorhinal area, precuneus, and parahippocampal gyrus. Differences in the remaining cortical and subcortical regions were less pronounced. The cognitively impaired groups showed larger and more spatially extensive mean absolute SHAP values than the cognitively normal groups. Higher values were observed across temporal and limbic regions, lateral and posterior cortical areas, and structures surrounding the ventricular system. Similar attribution patterns were observed among participants with NPI-Q scores of zero and those with positive scores. Because the model predicted continuous NPI-Q severity and the maps display absolute attributions, nonzero SHAP values among participants with scores of zero do not indicate that the model predicted the presence of NPS. Nevertheless, the stronger attribution patterns among cognitively impaired participants may partly reflect cognitive-status-related differences in brain morphology; therefore, these findings should be interpreted as patterns of model behavior rather than NPS-specific neuroanatomical effects.

Overall, the visual difference between cognitively normal participants and patients was more prominent than the difference between participants with zero and positive NPI-Q scores within either diagnostic group. These maps provide a descriptive comparison and do not establish statistically significant differences between the groups. Because absolute SHAP values were used, the maps show the magnitude of each regional contribution but not whether that contribution increased or decreased the predicted NPI-Q score.

To investigate whether NPS status was associated with different regional model-based contributions in cognitively normal individuals and patients along the AD continuum, we calculated Cohen’s d effect sizes using individualized regional NPSNetSHAP values. Within the cognitively normal and combined MCI/AD groups, participants without neuropsychiatric symptoms (NPI-Q = 0) were compared with those exhibiting symptoms (NPI-Q > 0). The two contrasts are presented side by side in **Figure 5** using a common effect-size scale. The cognitively normal comparison showed larger and more spatially extensive effects, particularly in frontal and parietal regions, with both positive and negative differences across the brain. In contrast, the MCI/AD comparison showed smaller, more diffusely distributed effects that were generally closer to zero.

**Figure 5.**
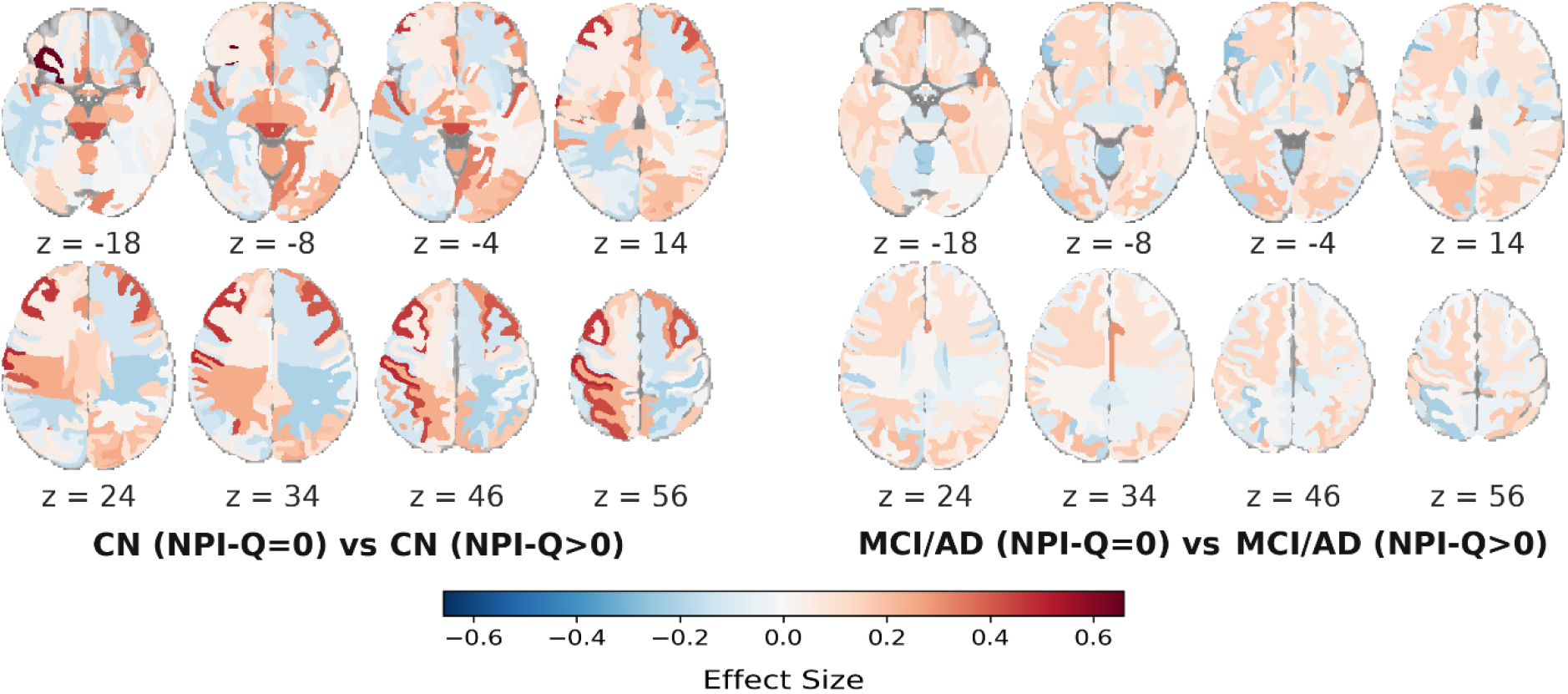
Neuropsychiatric symptom-related differences in individualized regional NPSNetSHAP values. Cohen’s d effect-size maps compare participants without neuropsychiatric symptoms (NPI-Q = 0) with those exhibiting symptoms (NPI-Q > 0) within the cognitively normal group (left) and the combined MCI/AD group (right).

## Discussion

This study examined whether regional sMRI features could predict NPS burden across clinically defined cognitive-status groups and whether the resulting predictions could be interpreted at the level of individual brain regions. Three findings stand out. First, all evaluated models produced significant predictions of NPI-Q total severity scores, with the NPSNet providing the strongest performance. The total severity score was selected as the prediction target because the study focused on overall NPS burden and because the total score has demonstrated stronger psychometric reliability than proposed NPI-Q subscales (40). Predicting specific symptom domains, such as apathy or psychosis, was beyond the scope of the present study but represents an important direction for future research. Second, the semi-simulated analysis showed that NPSNetSHAP recovered all ten predefined regional perturbations, compared with nine for NPSNetLIME and four for NPSNetLRP. Third, the individualized NPSNetSHAP patterns differed across cognitive-status groups, with the most extensive differences observed between cognitively normal participants and those with AD dementia. The principal methodological contribution of this study is the integration of whole-brain prediction, ground-truth validation of feature-attribution methods, and individualized regional interpretation within a single framework. Rather than interpreting an explainable model solely on the basis of its experimental output, we first tested whether its regional attributions recovered a known multivariate signal. Together, the findings suggest that predictions of NPS burden are informed by distributed and potentially nonlinear patterns of structural brain variation.

The NPSNet achieved the highest correlation between observed and predicted NPI-Q scores, followed by gradient boosting and SVR. The three nonlinear models therefore occupied the highest positions in the performance ranking, whereas Lasso, Elastic Net, and Ridge produced weaker correlations. Previous sMRI studies have demonstrated that deep-learning models can perform well in AD classification and the prediction of progression from MCI to AD (41). The present study extends this literature from diagnostic classification and disease-progression prediction to the continuous prediction of NPS burden. Within this task, the higher performance of the NPSNet and the other nonlinear models suggests that the imaging signal may involve nonlinear relationships or interactions among multiple regions that are not adequately represented by univariate methods or a simple linear combination of regional volumes.

The semi-simulated analysis provided useful validation of the explanation framework. NPSNetSHAP recovered all ten perturbed regions among its ten most influential features, whereas NPSNetLIME recovered nine and NPSNetLRP recovered four. The complete recovery by SHAP supports its use for the subsequent analysis of the experimental data and is consistent with its ability to quantify feature contributions while considering different feature combinations. Demonstrating that an attribution method can recover known ground-truth features provides greater confidence than applying it directly to observational data without validation. The lower recovery achieved by LIME and LRP also illustrates that different explanation methods can produce substantially different regional rankings even when applied to the same predictive model (20). Thus, the scientific contribution of the semi-simulated experiment lies not only in selecting SHAP but also in demonstrating a practical strategy for validating regional explanations before drawing neuroanatomical conclusions from them.

In the experimental dataset, the NPSNetSHAP maps showed that NPI-Q prediction depended on a spatially distributed pattern rather than on a single region. Contributions were evident in medial and inferior temporal regions, lateral temporal cortex, limbic structures, and regions surrounding the ventricular system. This pattern is broadly consistent with previous studies relating NPS to frontotemporal and limbic structural alterations. For example, reduced inferior temporal thickness has been associated with worsening apathy, whereas reduced supramarginal thickness has been associated with increasing hallucinations across the AD spectrum (13). Whole-brain analyses have also identified predominantly frontotemporal associations between NPS and brain morphology, with some associations differing between MCI and AD. More recent studies have reported prefrontal and temporal correlates of affective and vegetative symptoms (42) and distinct structural patterns associated with agitation, disinhibition, and irritability across cognitively normal, MCI, and AD groups. The present results therefore converge with earlier work at the level of temporal, limbic, and distributed association systems while extending it through simultaneous modeling of whole-brain features and participant-specific attribution maps. Differences in the exact regional patterns are expected because previous studies frequently examined individual symptoms or symptom factors, whereas the present model predicted total NPS burden.

The ventricular and periventricular findings may reflect broader neurodegenerative changes rather than a symptom-specific pathway. Ventricular expansion is closely related to surrounding tissue loss, while periventricular structural variation may capture diffuse disease effects not represented by a single cortical region.

The diagnostic-group analyses provided additional evidence that the model used different structural information across the AD continuum. Mean absolute SHAP values were more extensive in the AD group than in the cognitively normal and MCI groups. Similarly, the Cohen’s d effect size analysis showed the largest differences between cognitively normal and AD participants, whereas the CN vs. MCI comparison produced a more restricted pattern. The comparison involving the combined MCI and AD group showed an intermediate distribution. This ordering is consistent with greater separation of individualized attribution patterns at more advanced diagnostic stages. Previous morphometric studies have likewise found that associations between NPS and regional brain structure vary by diagnosis rather than remaining uniform across cognitively normal, MCI, and AD groups (15, 42, 43). For example, temporal associations with depression and anxiety and prefrontal associations with nighttime disturbance or apathy have been reported to differ across diagnostic comparisons (42), while hyperactive symptoms have shown distinct regional signatures in different diagnostic groups (43). The present study adds to this evidence by showing that diagnosis-related heterogeneity is also apparent in multivariate, individualized model explanations. However, because the data were cross-sectional, it cannot be concluded that these patterns evolved within individuals as their disease progressed. Longitudinal imaging and repeated NPI-Q assessments will be needed to determine whether changes in NPSNetSHAP profiles track the emergence or worsening of NPS.

Several features strengthen the present study. The analysis included a relatively large sample drawn from two cohorts, evaluated 145 whole-brain regional structural measures simultaneously, and directly compared the NPSNet with five conventional ML approaches. The repeated cross-validation design reduced dependence on a single data split, while the semi-simulated experiment provided known ground-truth regions for evaluating the explanation methods. Finally, the use of participant-level SHAP values moved the analysis beyond a single group-averaged feature ranking and allowed regional attribution patterns to be compared across diagnostic groups. Collectively, these features distinguish the proposed NPSNetSHAP framework from studies that focus only on predictive accuracy or apply an explanation method without first testing whether its attributions recover a known signal.

Combining ADNI and Knight-ADRC data increased the size and heterogeneity of the study population. This is a strength because models developed from a single cohort can capture acquisition characteristics or sampling patterns that do not generalize to other populations. The combined dataset incorporated variation in age, diagnosis, sMRI acquisition, and cohort composition. Harmonization was used to reduce the influence of these factors.

Some limitations of this study should be considered. First, the analysis focused on the total score from the Neuropsychiatric Inventory Questionnaire (NPI-Q). Although the total score provides a concise measure of overall neuropsychiatric symptom (NPS) burden and has demonstrated stronger psychometric reliability than proposed NPI-Q subscales (29), it combines symptom domains that may have different anatomical correlates. Domain-specific models of apathy, agitation, depression, hallucinations, and other symptoms may reveal more specific and clinically meaningful patterns. Second, despite the established psychometric properties of the NPI-Q total score, its ratings are based on caregiver observations and may remain influenced by differences in informant knowledge, contact, and reporting behavior. Third, the study used regional volumetric measures derived from structural magnetic resonance imaging (sMRI). Although this approach improves interpretability and reduces dimensionality, it may obscure finer subregional or spatially localized effects. Finally, although the ability to predict NPS burden using sMRI alone is promising, sMRI does not capture the full range of biological and clinical factors associated with NPS. Future multimodal models incorporating vascular injury, white-matter microstructure, functional connectivity, amyloid and tau biosignatures, medication exposure, psychosocial factors, and clinical or electronic health record data may improve predictive performance and clarify the mechanisms underlying NPS. Fifth, the semi-simulated experiment imposed perturbations within a predefined set of regions. Its findings therefore validate feature recovery under the specific conditions of that simulation rather than across all possible patterns of brain–behavior associations. Finally, Shapley additive explanation (SHAP) values are model-dependent. A different model architecture, feature representation, or training sample could produce a different attribution hierarchy. The moderate performance is also understandable given the nature of the outcome. The NPI-Q is a brief informant-based measure that combines several distinct behavioral and psychological domains into a single total score (5). Symptoms such as apathy, agitation, depression, anxiety, irritability, and hallucinations are unlikely to share a single anatomical substrate. They can also fluctuate over time and may be affected by medication, environmental circumstances, caregiver perception, physical health, and pre-existing psychiatric conditions. sMRI captures only one part of this broader clinical picture. Previous work has similarly shown that NPS vary across stages of cognitive impairment and are associated with heterogeneous structural changes (11, 42, 44–47). Accordingly, highly accurate prediction based on regional volumes alone would not necessarily be expected. Our results suggest moderate prediction accuracy, which could be due to the fact that brain-behavior relationships typically have small effect sizes when accurately assessed in a large sample (48). While this requires caution in terms of clinical applications, small or moderate effects can still provide critical clues to the underlying pathophysiology.

Future work should evaluate the proposed framework in independent and more clinically representative cohorts. Future longitudinal analyses could determine whether imaging-based predictions anticipate the onset or worsening of NPS. Modeling the individual NPI-Q domains, in addition to the total score, may help distinguish shared neurodegenerative effects from symptom-specific pathways and permit more direct comparison with previous regional studies. Investigating other modalities and variables, such as diffusion MRI, functional MRI, vascular imaging, amyloid and tau biosignatures, medication history, and relevant clinical measures will be important in the future. External validation, model calibration, stability testing of the regional explanations, and uncertainty estimates will be essential before the approach can be considered for clinical applications.

In conclusion, this study demonstrates that regional sMRI contains measurable information about NPS burden and that explainable AI can organize this information into individualized multivariate regional profiles. The explainable AI-based NPSNetSHAP framework extends previous neuroimaging work by combining whole-brain nonlinear prediction with ground-truth validation of regional explanations and diagnostic-group comparison. Its recovery of a known multivariate signals in semi-simulated data provides an empirical basis for interpreting the distributed contributions observed in the combined cohort of this study. These findings support explainable AI models as tools for decoding the whole-brain multivariate neuroanatomical correlates of NPS. At present, such models are best viewed as hypothesis-generating and complementary to clinical assessment rather than as replacements for direct evaluation of patients and caregivers. Although individual NPI-Q domains may have distinct neuroanatomical correlates, domain-specific modeling was beyond the scope of this initial framework and remains an important direction for future study.

## Data Availability

Data used in this study were obtained from the Alzheimer's Disease Neuroimaging Initiative (ADNI) and the Knight Alzheimer Disease Research Center (Knight-ADRC). Access is governed by the respective data repositories and may require registration, application, approval, and acceptance of applicable data-use agreements. Derived results generated in this study are available from the corresponding authors upon reasonable request, subject to applicable human-subject and data-transfer requirements.

## Code Availability Statement

The codes are publicly available at https://github.com/ganchand/NPSNetSHAP.

