## Supplementary Materials for "Explainable Deep Learning Reveals Distributed Neurodegeneration Signatures of Neuropsychiatric Symptoms Across the Alzheimer’s Continuum"

### S1. Experimental Data

Participants were drawn from the ADNI-2 and the Knight-ADRC. Inclusion in the present study required an available baseline T1-weighted sMRI scan and NPI-Q assessment. When more than one eligible NPI-Q assessment was available, the assessment obtained closest to the sMRI acquisition date was selected. The combined analytical sample included 1,756 participants.

#### S1.1. ADNI

Data used in this study were obtained from ADNI-2, a phase of the broader ADNI. ADNI is a multicenter, longitudinal public–private program established to investigate whether neuroimaging, biological markers, and clinical and neuropsychological assessments can be combined to characterize the progression of MCI and dementia due to AD. ADNI-2 extended the earlier ADNI phases by continuing the longitudinal follow-up of eligible participants and enrolling additional participants across the cognitive spectrum [1]. The ADNI-2 analytical sample included 668 participants (308 female; age range, 55.1–91.5 years; mean ± SD, 72.8 ± 7.3 years): 187 CN participants, 173 participants with EMCI, 152 with LMCI, 6 with a general MCI classification, and 150 with AD dementia. Newly enrolled ADNI-2 participants were generally required to be 55–90 years of age at screening, have a reliable study partner, be proficient in English or Spanish, and be willing and able to complete longitudinal clinical, cognitive, biomarker, and imaging assessments. Participants continuing from ADNI-1 or ADNI-GO had originally been enrolled according to the applicable protocols for those phases and remained eligible for longitudinal follow-up in ADNI-2. For newly enrolled participants, diagnostic classification was based on the Mini-Mental State Examination (MMSE), global Clinical Dementia Rating (CDR), education-adjusted delayed-recall performance on the Wechsler Memory Scale–Revised Logical Memory II subtest, subjective memory concerns, functional status, and clinical assessment. CN participants had an MMSE score of 24–30, a global CDR score of 0, and delayed-recall performance within the education-adjusted normal range. Participants classified as EMCI or LMCI had an MMSE score of 24–30, a subjective memory concern reported by the participant, study partner, or clinician, a global CDR score of 0.5, generally preserved activities of daily living, and no diagnosis of dementia. EMCI was defined by Logical Memory II delayed-recall scores of 9–11 for participants with at least 16 years of education, 5–9 for those with 8–15 years, and 3–6 for those with 0–7 years. The corresponding LMCI thresholds were ≤8, ≤4, and ≤2, respectively. Participants classified as AD had an MMSE score of 20–26, a global CDR score of 0.5 or 1.0, and met established NINCDS/ADRDA criteria for probable AD. Enrollment was also subject to protocol-specific criteria concerning depression, medical stability, and the use of psychoactive medications [1]. For the present group analyses, participants classified as EMCI, LMCI, or general MCI were combined into a single MCI group.

#### S1.2. Knight-ADRC

The Knight-ADRC at Washington University in St. Louis maintains a longitudinal cohort of community-dwelling volunteers spanning cognitively normal aging, MCI, and AD. Participants undergo standardized clinical and neuropsychological assessments, neuroimaging, biomarker collection, and longitudinal follow-up, providing a deeply phenotyped resource for the investigation of AD and related dementias. The Knight-ADRC analytical sample included 1,088 participants (605 female; age range, 45.2–91.4 years; mean ± SD, 70.4 ± 8.5 years): 920 CN participants and 168 participants classified within a combined MCI/AD group. Clinical status was established through standardized clinical and cognitive assessments, including interviews with the participant and a knowledgeable collateral source, determination of the global CDR score, and the clinician's etiologic assessment. A global CDR score of 0 indicated no cognitive impairment, whereas scores of 0.5, 1, 2, and 3 represented increasing levels of cognitive and functional impairment. Participants with a global CDR score of 0 were classified as CN. Participants with a global CDR score of 0.5 or higher and a clinical presentation attributed to MCI or AD were included in the symptomatic group. Individuals whose primary diagnosis reflected a non-AD cause of cognitive impairment, including vascular dementia, dementia with Lewy bodies, frontotemporal dementia, or Parkinson's disease, were excluded. Participants with medical or neurological conditions that prevented reliable longitudinal participation were also excluded. All participants or their legally authorized representatives provided informed consent under protocols approved by the relevant institutional review board.

Because separate MCI and AD labels were not available in the Knight-ADRC dataset used in the present study, symptomatic participants from this cohort were retained as a combined MCI/AD group. Thus, the cohort-specific diagnostic information was preserved during harmonization: ADNI-2 participants classified as EMCI, LMCI, or general MCI were combined into an MCI group, whereas symptomatic Knight-ADRC participants remained in the combined MCI/AD category.

### S2. Semi-simulated data generation

We generated a semi-simulated dataset to determine whether the NPSNetSHAP method could recover a known multivariate relationship between regional brain volumes and NPI-Q scores. The dataset was based on structural MRI data from 1,107 cognitively normal participants in the combined ADNI and Knight-ADRC cohorts. Using cognitively normal participants provided a relatively clean baseline in which the synthetically introduced changes could be interpreted as disease-like patterns.

For each participant, volumetric measures were obtained from 145 regions of interest segmented using MUSE. The regional measures were harmonized to account for cohort, imaging site, age, sex, and total intracranial volume. Each harmonized regional volume was then divided by the sum of all regional volumes for the same participant. This was the same preprocessing procedure used for the experimental data.

Ten medial temporal and limbic regions were selected for synthetic perturbation. These regions were chosen to create a predefined multivariate pattern that could serve as the ground truth for evaluating the feature-attribution methods.

Synthetic neurodegenerative patterns were introduced in 50% of the participants. For each selected participant, the volume of every ground-truth region was reduced by a randomly sampled value ranging from 10% to 30%. The corresponding NPI-Q score was increased in proportion to the mean fractional volume reduction across the perturbed regions. This increase was scaled to the valid NPI-Q range, and Gaussian noise with a standard deviation of one was added. The resulting NPI-Q scores were rounded to the nearest integer. This procedure imposed an inverse relationship between regional brain integrity and neuropsychiatric symptom burden. The participant-specific perturbations preserved variation in the strength of this relationship and prevented all pseudo-patient observations from having the same pattern.

Regions outside the predefined ground-truth set were not directly modified. The semi-simulated dataset therefore retained the covariance structure and interindividual variation of the original imaging data while containing a known multivariate signal.

The semi-simulated data were analyzed using the same cross-validation and model-training procedures used for the experimental data. Prediction performance was evaluated using Spearman's correlation between the simulated NPI-Q scores and their corresponding held-out predictions. The NPSNet showed a strong correlation between the observed and predicted scores ($\rho = 0.93,\ p < 2.2 \times 10^{-308}$).

The known perturbation pattern was also used to evaluate explanation accuracy. For each attribution method, the brain regions were ranked according to their mean absolute attribution values. Performance was defined as the number of ground-truth regions recovered among the ten highest-ranked features.

### S3. Cross-validated prediction performance on semi-simulated data

Model performance was assessed by 10-fold cross-validation over the 2,000 semi-simulated subjects. Each subject was scored exclusively by the model for which it was held out, so the fold-wise statistics reported in Table X constitute out-of-sample estimates throughout. Agreement between predicted and ground-truth NPI-Q was quantified by Spearman's rank correlation coefficient (ρ) reported on the native NPI-Q scale (0-20). The network recovered the simulated ROI-symptom mapping with high fidelity and low between-fold variance and all fold-wise correlations were significant at $p < 10^{-78}$. Pooling all 2,000 held-out predictions into a single distribution yielded a marginally attenuated coefficient ($\rho = 0.927$), as aggregation across folds superimposes the residual calibration differences between independently trained models.

***Table S3.1.*** Fold-wise prediction performance on the semi-simulated cohort. Each fold comprises 200 held-out subjects scored by the model trained on the remaining nine folds. $\rho$, Spearman rank correlation between predicted and ground-truth NPI-Q.

| **Fold** | $n$ | $\rho$ | $p$ |
|---|---|---|---|
| 1 | 200 | 0.963 | $1.95 \times 10^{-114}$ |
| 2 | 200 | 0.949 | $2.44 \times 10^{-101}$ |
| 3 | 200 | 0.942 | $1.58 \times 10^{-95}$ |
| 4 | 200 | 0.952 | $8.59 \times 10^{-104}$ |
| 5 | 200 | 0.948 | $1.04 \times 10^{-100}$ |
| 6 | 200 | 0.912 | $2.05 \times 10^{-78}$ |
| 7 | 200 | 0.941 | $4.39 \times 10^{-95}$ |
| 8 | 200 | 0.935 | $6.46 \times 10^{-91}$ |
| 9 | 200 | 0.924 | $7.33 \times 10^{-85}$ |
| 10 | 200 | 0.948 | $1.65 \times 10^{-100}$ |

### S4. NPSNet hyperparameter tuning

Hyperparameter tuning was performed to identify a NPSNet configuration that provided stable NPI-Q prediction while limiting overfitting. Candidate configurations were evaluated using the semi-simulated dataset. The primary performance measure was Spearman's correlation between actual and predicted NPI-Q scores in the held-out test data. Mean absolute error was used as a complementary measure of prediction error if it was retained in the final analysis.

Each hyperparameter configuration was evaluated using ten-fold cross-validation repeated across five independent runs. Within each fold, 90% of the observations were used for model development and 10% were reserved for testing. A further 5% of the training observations were used as an internal validation set. This validation set was used to monitor model training and apply early stopping. The independent test fold was not used for hyperparameter selection, early stopping or model fitting.

The NPSNet model consisted of fully connected hidden layers with rectified linear unit activation. Batch normalization and dropout were applied after each hidden layer, and the output layer contained a single unit with linear activation. Network weights were initialized using a random normal distribution. Training was performed using the Adam optimizer and mean absolute error loss. Training was allowed to continue for a maximum of 1000 epochs, with early stopping applied after 50 consecutive epochs without improvement in validation loss. The weights producing the lowest validation loss were restored before test-set evaluation.

Hyperparameters should be selected using only the training and validation observations within each cross-validation fold. The final configuration should be determined from performance averaged across the repeated runs rather than from a single random partition.

#### S4.1. Learning-rate selection

The effect of the learning rate was evaluated while the remaining hyperparameters were held at their default values. The selected learning rate should correspond to the configuration providing the strongest held-out Spearman correlation together with an acceptably low prediction error.

***Table S4.1.*** *NPSNet performance across candidate learning rates.*

| Learning rate | Spearman ρ | p-value | Test MAE |
|---|---|---|---|
| 0.001 | 0.217 | $7.69 \times 10^{-6}$ | 3.458 |
| 0.005 | 0.134 | $3.55 \times 10^{-5}$ | 3.124 |

| **Learning rate** | **Spearman ρ** | **p-value** | **Test MAE** |
|---|---|---|---|
| 0.010 | 0.110 | $7.56 \times 10^{-1}$ | 3.166 |
| 0.050 | 0.500 | $7.02 \times 10^{-11}$ | 2.290 |
| $\mathbf{0.100}$ | $\mathbf{0.605}$ | $\mathbf{5.44 \times 10^{-27}}$ | $\mathbf{1.815}$ |

### S4.2. Batch-size selection

Candidate batch sizes were evaluated using the learning rate selected in Section S2.1. All remaining model settings were held constant. The selected batch size was the configuration providing the best average performance across the repeated cross-validation runs.

***Table S4.2.*** *NPSNet performance across candidate batch sizes.*

| **Batch size** | **Spearman $\rho$** | **$p$-value** | **Test MAE** |
|---|---|---|---|
| $\mathbf{16}$ | $\mathbf{0.766}$ | $\mathbf{3.65 \times 10^{-35}}$ | $\mathbf{1.040}$ |
| 32 | 0.796 | $4.37 \times 10^{-36}$ | 0.880 |
| 64 | 0.567 | $6.42 \times 10^{-27}$ | 1.956 |
| 128 | 0.159 | $2.34 \times 10^{-4}$ | 3.020 |

### S4.3. Network-architecture selection

Network depth and the number of units in each hidden layer were evaluated to identify an architecture sufficiently flexible to capture nonlinear associations without excessive complexity. Candidate architectures should be listed exactly as implemented.

***Table S4.3.*** *NPSNet performance across candidate network architectures.*

| **Hidden layers and units** | **Spearman $\rho$** | **$p$-value** | **Test MAE** |
|---|---|---|---|
| $\mathbf{40 - 30 - 20 - 10}$ | $\mathbf{0.796}$ | $\mathbf{4.37 \times 10^{-36}}$ | $\mathbf{0.880}$ |
| $40 - 25 - 10$ | 0.792 | $1.31 \times 10^{-37}$ | 1.003 |
| $64 - 32 - 16$ | 0.770 | $4.66 \times 10^{-34}$ | 1.114 |
| $32 - 16$ | 0.792 | $1.17 \times 10^{-34}$ | 1.239 |

The selected learning rate, batch size, architecture, dropout rate, optimizer and early-stopping settings were used for all analyses reported in the main manuscript.

### S5. Comparison of feature-attribution methods

Three feature-attribution methods were compared to determine which approach most accurately recovered the predefined regional pattern in the semi-simulated dataset: SHAP, LIME and LRP. Each method was applied to the trained NPSNet, resulting in the NPSNetSHAP, NPSNetLIME and NPSNetLRP frameworks.

#### S5.1. SHAP

SHAP is a state-of-the-art approach for the interpretability and explainability of complex predictive models, providing a global perspective by revealing the overall significance of each input feature towards the prediction, hence explaining the black box models like DL models. It satisfies all three essential properties concerning interpretability, explainability, and accuracy of the black box AI model, and those properties are local accuracy, missingness, and consistency. Local accuracy ensures that the SHAP explanation model faithfully reproduces the original model's prediction for a specific instance by guaranteeing that the sum of the SHAP values across all input features equals the output of the original model. When all features are set to their baseline (reference) values, the explanation model's prediction matches the model's expected prediction under the same condition. Missingness guarantees that the absence of certain features does not affect the interpretation, while consistency ensures that feature importance remains unchanged unless a feature's contribution alters.

SHAP quantifies the significance of an input feature for a model's output through its SHAP value, which is derived from the Shapley value in cooperative game theory. Through Shapley value players in a game are assigned fair values based on their contribution to the game's output. The SHAP value for an input feature ($i$) is computed through the given formula:

$$\phi_i(f, x) = \sum_{z' \subseteq x'} \frac{(|z'| - 1)!\,(M - |z'|)!}{M!} [f_x(z') - f_x(z' \setminus i)],$$

where, $f$ denotes the trained model, ($x = [x_1, x_2, \ldots, x_M]$) represents the input features, and $x'$ refers to simplified binary input vectors indicating which features are included (1) or excluded (0). Each subset $z'$ of $x'$ reflects a specific combination of present or absent features, and the term $f_x(z')$ is the expected model output when only the features in $z'$ are known. The difference $f_x(z') - f_x(z' \setminus i)$ captures the marginal effect of including feature $i$, and the weighting factor

$\frac{(|z'|-1)!(M-|z'|)!}{M!}$ ensures fair contribution from each subset, derived from cooperative game theory principles.

### S5.2. LIME

LIME explains an individual prediction by generating perturbed observations around the target case and fitting an interpretable surrogate model to approximate the NPSNet locally. The explanation model is obtained by minimizing

$$\xi(x) = \text{argmin}_{g \in G}[L(f, g, \pi_x) + \Omega(g)],$$

where $f$ is the original NPSNet, $g$ is an interpretable surrogate model, $L$ measures the fidelity of the surrogate within the local neighborhood defined by $\pi_x$, and $\Omega(g)$ penalizes model complexity.

LIME was implemented in regression mode using the tabular explanation interface. The feature-selection setting, number of simulated samples and number of features returned for each explanation are as follows:

- Feature-selection method: highest weights
- Number of perturbed samples: 5000
- Number of reported features: 145

### S5.3. LRP

LRP assigns relevance scores by propagating the NPSNet output backward through the network while approximately conserving the total relevance across layers. Under the epsilon rule, relevance assigned to input feature $i$ is calculated as

$$R_i^{(l)} = \sum_j \frac{z_{ij}}{\sum_{i'} z_{i'j} + \epsilon \text{sign}\left(\sum_{i'} z_{i'j}\right)} R_j^{(l+1)},$$

where $z_{ij}$ represents the contribution from neuron $i$ to neuron $j$, $R_j$ is the relevance assigned to neuron $j$, and $\epsilon$ is a stabilizing constant.

### S5.4. Ground-truth recovery

The three methods were evaluated according to their ability to identify the ten perturbed regions. For each method, attribution values were converted to absolute values and averaged across participants and repeated model runs. The regions were then ranked according to their mean absolute attribution values.

NPSNetSHAP recovered all ten (100%) ground-truth regions among its ten highest-ranked features. NPSNetLIME recovered nine (90%), missing the right parahippocampal gyrus. NPSNetLRP recovered four (40%): the bilateral amygdala, left entorhinal area and left precuneus while the remaining six NPSNetLRP features were not perturbed.

***Table S5.1.*** *Recovery of the ground-truth regions by the feature-attribution methods.*

| **Perturbed region** | **Top 10 NPSNetSHAP** | **Top 10 NPSNetLIME** | **Top 10 NPSNetLRP** |
|---|---|---|---|
| Right hippocampus | Yes | Yes | No |
| Left hippocampus | Yes | Yes | No |
| Right amygdala | Yes | Yes | Yes |
| Left amygdala | Yes | Yes | Yes |
| Right entorhinal area | Yes | Yes | No |
| Left entorhinal area | Yes | Yes | Yes |
| Right precuneus | Yes | Yes | No |
| Left precuneus | Yes | Yes | Yes |
| Right hippocampal | Yes | Yes | No |
| Right parahippocampal gyrus | Yes | No | No |
| **Recovery rate** | **100%** | **90%** | **40%** |

These findings indicate that SHAP most accurately recovered the known regional pattern in the present semi-simulated experiment. NPSNetSHAP was therefore selected for interpreting NPI-Q predictions in the experimental ADNI and Knight-ADRC dataset.

### S6. Regional NPSNetSHAP patterns according to diagnostic and NPI-Q status

In this section, we provide the results for participants with MCI or AD separately. Mean absolute regional NPSNetSHAP values were calculated separately for each group and projected onto a standard MRI template. The same color scale was used for all maps to allow direct visual comparison.

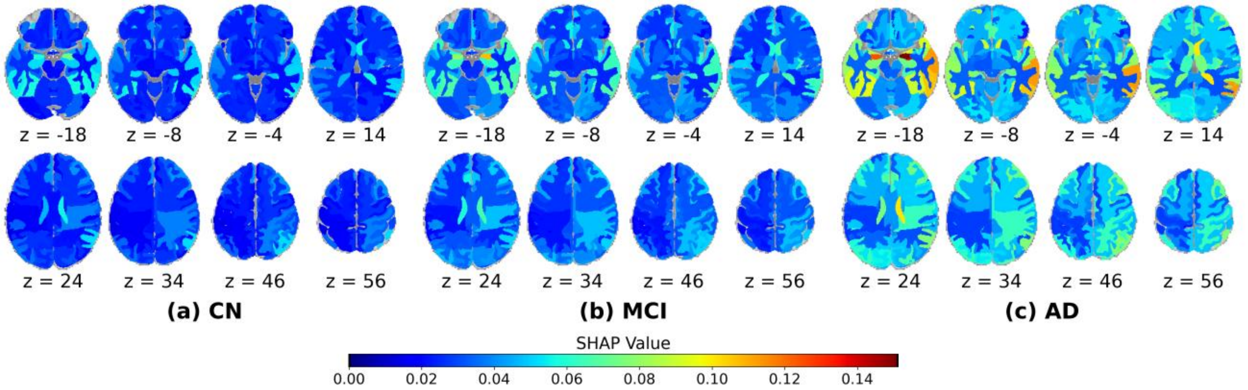


***Figure S6.1.*** *Regional NPSNetSHAP patterns according to diagnostic groups.*

### S7. Separate Comparisons for MCI and AD

The comparison between the cognitively normal and AD groups produced the largest and most widespread effects. The strongest group differences were concentrated in bilateral medial and inferior temporal regions, with additional effects involving lateral temporal cortex and ventricular or periventricular structures. The comparison between cognitively normal participants and those with MCI showed a similar but more restricted anatomical pattern. Effect sizes were generally smaller than those observed in the CN-AD comparison and were concentrated primarily in medial and lateral temporal regions. The comparison between cognitively normal participants and the combined MCI and AD group produced an intermediate pattern. Differences remained prominent in medial temporal and temporal neocortical regions and extended to several ventricular and posterior cortical structures. Across the three comparisons, positive effects were more extensive than negative effects, while negative effects were smaller and spatially limited. Overall, the effect-size maps show that the individualized structural contributions underlying NPI-Q prediction became more distinct from the cognitively normal pattern in clinically affected groups, with the clearest separation observed for AD.

### REFERENCE

[1] https://adni.loni.usc.edu/wp-content/themes/freshnews-dev-v2/documents/clinical/ADNI-2_Protocol.pdf
